# Baseline gut microbiome and metabolites are correlated with alcohol consumption in a zonisamide clinical trial of heavy drinking alcoholic civilians

**DOI:** 10.1101/2024.04.02.24305199

**Authors:** Liv R. Dedon, Hanshu Yuan, Jinhua Chi, Haiwei Gu, Albert J. Arias, Jonathan M. Covault, Yanjiao Zhou

## Abstract

Development and severity of alcohol use disorder (AUD) has been linked to variations in gut microbiota and their associated metabolites in both animal and human studies. However, the involvement of the gut microbiome in alcohol consumption of individuals with AUD undergoing treatment remains unclear. To address this, stool samples (n=48) were collected at screening (baseline) and trial completion from a single site of a multi-site double-blind, placebo-controlled trial of Zonisamide in individuals with AUD. Alcohol consumption, gamma-glutamyl transferase (GGT), and phosphatidylethanol (PEth)levels were measured both at baseline and endpoint of 16-week trial period. Fecal microbiome was analyzed *via* 16S rRNA sequencing and metabolome *via* untargeted LC-MS. Both sex (p = 0.003) and psychotropic medication usage (p = 0.025) are associated with baseline microbiome composition. The relative abundance of 12 genera at baseline was correlated with percent drinking reduction, baseline and endpoint alcohol consumption, and changes in GGT and PeTH over the course of treatment (p.adj < 0.05). Overall microbiome community structure at baseline differed between high and low responders (67-100% and 0-33% drinking reduction, respectively; p = 0.03). A positive relationship between baseline fecal GABA levels and percent drinking reduction (R=0.43, p < 0.05) was identified by microbiome function prediction and confirmed by ELISA and metabolomics. Predicted microbiome function and metabolomics analysis have found that tryptophan metabolic pathways are over-represented in low responders. These findings highlight importance of baseline microbiome and metabolites in alcohol consumption in AUD patients undergoing zonisamide treatment.

## Introduction

Alcohol use disorder (AUD) persists as a major global public health challenge. AUD causes a high disease burden accounting for over 5% of annual deaths globally.^1,2^ Chronic heavy alcohol consumption has been associated with higher risks of developing other chronic health conditions such as cancer, dementia, digestive disorders, and liver disease.^3–6^ Currently, three are three FDA-approved drugs to treat AUD, but their efficacy is limited. Several repurposed agents such as Zonisamide have shown to reduce alcohol consumption and craving in clinical trials. ^7–9^ However, the degree of reduction in alcohol consumption varies greatly among patients treated with the drug or placebo even after controlling for all available factors. The act of taking placebo medication and meeting regularly with a healthcare provider alone can significantly influence reduction in alcohol consumption.^10^ This differential response to treatment is common in many clinical trials including AUD, but underlying causes remain elusive.

Recently, there has been growing interest in the bidirectional relationship between the development, progression, and treatment of AUD and the gut microbiome *via* the gut-brain axis. Both acute and chronic alcohol consumption are linked to a shift in microbiome composition favoring an overrepresentation of proinflammatory microbes and an underrepresentation of short-chain fatty acid-producing microbes.^11–17^ During periods of heavy alcohol consumption, the production of short-chain fatty acids (SFCAs) are significantly decreased.^11,16,18^ These microbially-produced SCFAs influence various biological processes such as depression, anxiety, and craving,^19–23^ which are common comorbidities of AUD. Further, the administration of SCFAs has been shown to reduce drinking behavior in rodent models.^23–26^ The microbiome and its metabolites are not only responsive to alcohol consumption but also able to influence drinking behavior. The AUD-associated gut microbiome has been linked to increased alcohol consumption,^27,28^ depression, anxiety, alcohol craving, and possibility of relapse.^7,24–28^ Manipulation of the microbiome has been shown to modify alcohol consumption in both animal models and humans. Antibiotic treatment increased binge drinking in mice^29^ but decreased binge drinking in rats.^30^ In rats, fecal microbiota transplantation from alcohol consuming donor rats increased voluntary drinking in naïve recipients, but this increase was subsequently reversed by antibiotic treatment.^27^ Fecal microbiota transplantation from healthy donors without AUD to patients with AUD has proven to be a promising route for treatment of AUD and AUD-associated health problems.^23,29^

The baseline gut microbiome has been found to play an important role in response to cancer therapy,^31^ nutritional intervention^32^ and efficacity of SARS-CoV-2 vaccines.^33,34^ However, whether the importance of baseline gut microbiome in alcohol drinking behavior in AUD patients undergoing treatment has not been investigated. We hypothesize that the gut microbiome and metabolites at baseline before treatment intervention are important factors influencing alcohol drinking behavior. To test this hypothesis, we leverage a randomized, placebo-controlled, double-blind clinical trial (ClinicalTrials.gov identifier: NCT02900352) recently completely by our research team testing effect of Zonisamide treatment of heavy drinking alcoholic civilians. Using 16S rRNA gene sequencing and untargeted metabolomics, we show reduction in alcohol consumption between baseline and endpoint visits of the clinical trial is significantly correlated with composition of the baseline microbiome and levels of specific gut metabolite such as GABA. Patients who achieved a high level of drinking reduction during the trial (high responders) had a distinct gut microbiome profile and specific gut metabolic signatures related to tryptophan metabolism, compared to patients who achieved a lower level of drinking reduction (low responders). Our findings highlight importance of baseline gut microbiome and metabolites in alcohol consumption and the potential for development of microbiome signature to predict treatment response for AUD.

## Results

### Study participants

The demographic and clinical characteristics and relevant alcohol-related metadata of the 48 patients with AUD included in the analysis are reported in Table 1. No significant differences in metadata distributions were observed between the zonisamide and placebo treatment groups. Participants were overall 44% female. Participant age ranged from 23 to 70 years with a median age of 56. More than half of the participants identified as white/non-Hispanic. While a higher percentage of participants in the zonisamide group dropped out of the study, there was a comparable number of completing participants in both treatment groups. In total, 48 participants provided baseline stool samples. 32 participants completed the study and have endpoint alcohol consumption and drinking reduction results. Of the completing participants, 21 participants provided an endpoint stool sample.

**Table 1:** Demographic and clinical metadata for participants in our study.

| Variable | Overall, N = 48<sup>1</sup> | Placebo, N = 24 | Treated, N = 24 | p-value |
| --- | --- | --- | --- | --- |
| **Completed study, n (%)** |  |  |  | 0.122 |
| Completed | 32 (67%) | 19 (79%) | 13 (54%) |  |
| Dropped out | 16 (33%) | 5 (21%) | 11 (46%) |  |
| **Sex, n (%)** |  |  |  | 0.22 |
| Male | 27 (56%) | 11 (46%) | 16 (67%) |  |
| Female | 21 (44%) | 13 (54%) | 8 (33%) |  |
| **Age, Median (IQR)** | 56 (50, 63) | 54 (49, 59) | 61 (52, 64) | 0.103 |
| **Race, n (%)** |  |  |  | 0.22 |
| White/Hispanic | 4 (8.3%) | 0 (0%) | 4 (17%) |  |
| White/Non-Hispanic | 39 (81%) | 21 (88%) | 18 (75%) |  |
| Black/Hispanic | 1 (2.1%) | 1 (4.2%) | 0 (0%) |  |
| Black/Non-Hispanic | 4 (8.3%) | 2 (8.3%) | 2 (8.3%) |  |
| **BMI, Median (IQR)** | 28.7 (25.3, 32.5) | 29.6 (26.5, 33.2) | 27.9 (23.7, 31.6) | 0.3 |
| **Prescribed Psychotropic Medication, n (%)** | 21 (44%) | 11 (46%) | 10 (42%) | >0.9 |
| **Smoker, n (%)** | 8 (17%) | 4 (17%) | 4 (17%) | >0.9 |
| **PEth, Median (IQR)** | 388 (145, 550) | 350 (129, 609) | 418 (225, 489) | 0.8 |
| **GGT, Median (IQR)** | 41 (28, 84) | 44 (29, 87) | 40 (26, 57) | 0.6 |
| **Total drinks in the 90 days prior to screening, Median (IQR)** | 457 (302, 557) | 394 (269, 590) | 483 (358, 540) | 0.6 |
| **Average drinks per week at screening, Median (IQR)** | 36 (24, 43) | 31 (21, 46) | 38 (28, 42) | 0.6 |
| **Average drinks per week during last 4 weeks of study, Median (IQR)** | 15 (7, 28) | 10 (4, 26) | 20 (14, 28) | 0.2 |
| **Percent drinking reduction, Median (IQR)** | 57 (34, 87) | 61 (36, 91) | 52 (29, 62) | 0.2 |

<sup>1</sup>n (%); Median (IQR)

From the entirety of the multi-site trial, mean drinks per day was significantly affected by zonisamide treatment (F_(3,1655)_ = 4.47, p=0.035) resulting in the placebo group reporting 0.72 (95% CI 0.54-0.93) more drinks per day than the zonisamide group.^35^ Zonisamide treatment was found to be more effective in male participants with the zonisamide group reporting 43% fewer drinks per week than the placebo group.

Female participants did not show a significant difference in drinks per week between treatment group. From the participants that provided stool samples used in this study, treatment (placebo group vs zonisamide group) was not correlated with percent drinking reduction from average drinks per week at baseline to average drinks per week over the last four weeks of the study reported at study endpoint (ANOVA, p = 0.3). Further, no significant correlation was found between percent drinking reduction and any other study metadata except average drinks per week, which is used to calculate the percent drinking reduction. Thus, treatment was not considered a confounding variable in the subsequent microbiome and metabolomic analysis of stool samples.

### Baseline microbiome composition is sex- and medication-linked

We first examined whether any demographic factors are associated with the baseline microbiome. Genus-level composition did not vary between treatment groups or across the study duration (Supporting Figure 1). Alpha and beta diversity show no significant difference between groups (p > 0.1 for all comparisons, Supporting Figures 2 & 3). Clear separation is observed in a PCoA plot of male and female microbiome beta diversity (Fig 1a, 1c). This difference is confirmed by PERMANOVA analysis (p = 0.003, Figure 1a). Linear discriminant analysis effect size analysis (LEfSe)^36^ detected 13 genera showing significant differences in relative abundance between female and male participants (Figure 1b). *Bacteroides* (LDA Score = 4.46, p = 0.013) and *Blautia* (LDA Score = 4.46, p = 0.033) were the most overrepresented genera in female participants at baseline. Male participant microbiomes showed an overrepresentation of *Enterococcus* (LDA Score = 3.81, p = 0.011) and *Veillonella* (LDA Score = 3.48, p = 0.025).

**Figure 1:**
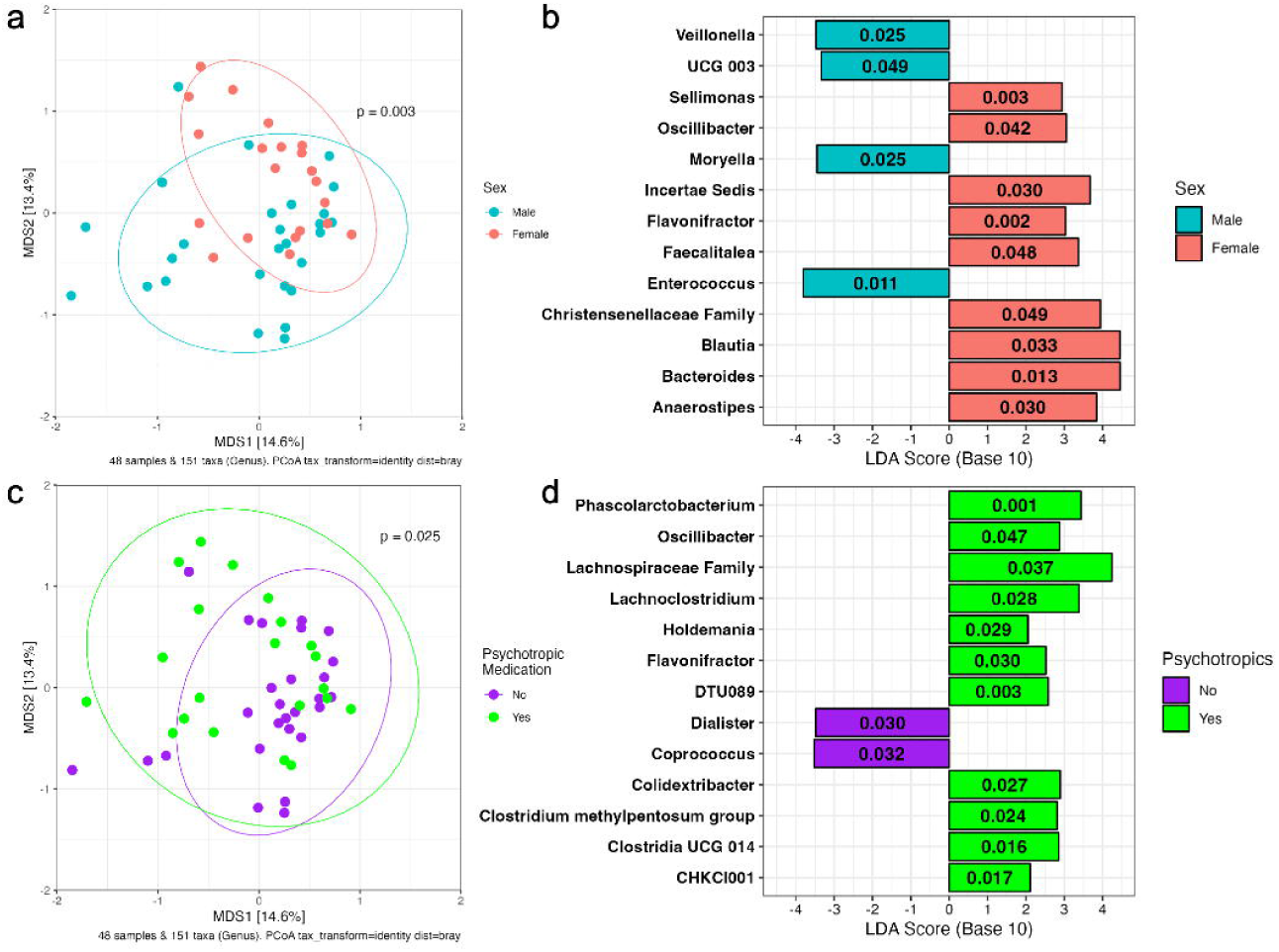
Associations between baseline gut microbiome and clinical characteristics of the participants in the study. (a) Baseline microbiome composition varies based on sex with male and female participants clustering separately in the PCoA plot (p = 0.003 by PERMANOVA based on Bray-Curtis dissimilarity). (b) 13 genera overrepresented in male or female participant microbiomes. (c) Baseline microbiome composition also varies based on psychotropic medication usage (PERMANOVA, p = 0.025). (d) 13 genera overrepresented in psychotropic medication users and non-users.

**Figure 2:**
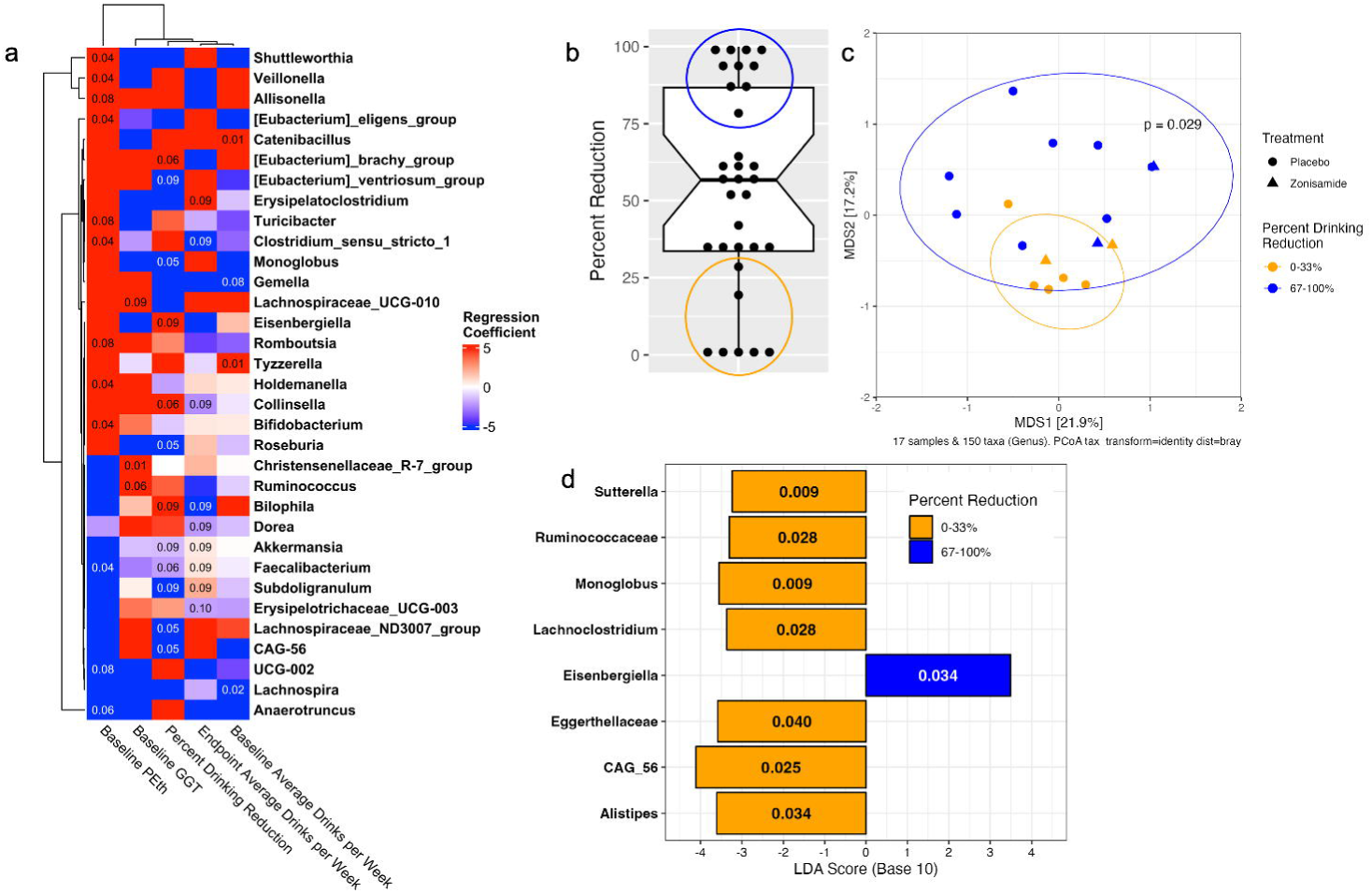
Baseline gut microbiome composition is associated with drinking reduction. (a) Genus-level relative abundance is correlated with percent drinking reduction over the study duration, baseline and endpoint alcohol consumption, and alcohol-related biomarkers PEth & GGT. Colors of the heatmap represent correlation coefficients derived from the multiple linear regression analysis. An adjusted p value is denoted in the cell of the heatmap if the adjusted p value is less than 0.1 for a given correlation analysis. (b) Percent drinking reduction is largely varied across participants and can be broken into tertiles. (c) The highest (percent drinking reduction 67-100%) and lowest (percent drinking reduction 0-33%) tertiles cluster separately in the PCoA plot (PERMANOVA p = 0.029). (d) 8 genera significantly overrepresented in the high- and low-responder groups.

We also found the microbiome in psychotropic medication users clustered separately from non-users in a PCoA plot. Further, PERMANOVA analysis confirmed overall microbiome difference between these two groups (p = 0.025, Figure 1c). LEfSe analysis revealed that psychotropic medication promotes over-representation of 11 genera (Figure 1d). Only 2 genera were over-represented in psychotropic medication non-users.

### Baseline microbiome is associated with alcohol consumption

Results from the 32 participants that completed the study were used to explore correlation between baseline microbiome and alcohol consumption at both baseline and endpoint as well as drinking reduction over the course of the study after controlling for sex and psychotropic medication and adjusting for multiple comparisons (Figure 2a). *Eubacterium brachy* group (p.adj = 0.05), *Bilophila* (p.adj = 0.09), and *Collinsella* (p.adj = 0.06) were positively correlated with percent drinking reduction. *Monoglobus* (p.adj = 0.05), *Roseburia* (p.adj = 0.05), *Lachnospiraceae* ND3007 group (p.adj = 0.05), *Subduogranulum* (p. adj = 0.09), *Akkermansia* (p.adj = 0.08), and *Faecalibaterium* (p.adj = 0.06) were all negatively correlated with percent drinking reduction.

In addition to self-reported alcohol consumption, serum biomarkers gamma-glutamyl transferase (GGT) and phosphatidylethanol (PEth) were also measured at baseline and endpoint visits (Fig 2a). Correlations between baseline microbiome and alcohol consumption-related biomarkers are largely unique from self-reported endpoint average drinks per week and percent drinking reduction. *Faecalibacterium* (p.adj = 0.04) was negatively associated with baseline PEth and percent drinking reduction (p.adj = 0.06), but positively correlated with endpoint average drinks per week (p.adj = 0.09). *Clostridium sensu stricto* 1was positively correlated with baseline PEth (p.adj = 0.04) and negatively associated with endpoint average drinks per week (p.adj = 0.09). *Ruminococcus* was positively correlated with baseline GGT (p.adj = 0.06). It is of note that neither serum GGT and PEth levels nor baseline drinks per week were significantly correlated with percent drinking reduction at this trial site (Fisher’s exact test, p >0.05 for all comparisons).

### Baseline microbiome composition differs between high and low responders

Percent drinking reduction showed a high inter-subject variation (Figure 2b). To have a better understanding the involvement of the microbiome in the “high vs low responders”, the spread of percent drinking reduction across patients was broken into tertiles. Of the 32 completing participants, 10 had a “high response” classified henceforth as percent drinking reduction of 67-100% (Figure 2b, blue circle). Seven participants had a “low response” classified henceforth as percent drinking reduction of 0-33% (Figure 2b, orange circle). High responders clustered separately from low responders from PCoA analysis (Figure 2c). Baseline microbiome composition varied significantly between the high and low responders from PERMANOVA (p= 0.029). Alpha diversity was consistent between high and low responders (Supporting Figure 4). LefSE identified 7 genera that were significantly overrepresented in low responders and 1 genus that was significantly overrepresented in high responders (Figure 2d). Many of the genera identified by LefSE have previously been associated with alcohol consumption. CAG-56 is significantly overrepresented in low responders (p.adj = 0.025) and shows a negative association trend with percent drinking reduction (p.adj = 0.05). *Eisenbergiella* is overrepresented in high responder baseline microbiome (p.adj = 0.034) and positively associated with percent drinking reduction (p.adj = 0.09).

### Baseline GABA levels in the stool are correlated with drinking reduction

The gut microbiome has been shown to modulate production of many neurotransmitters involved in AUD. To understand the potential involvement of neuroactive potential of the gut microbiome in drinking response, we predicted neuroactive compound production or degradation process based on the gut microbiome using the gut-brain module (GBM) analysis. ^37–39^ We found a positive linear correlation between percent drinking reduction and potential for γ-aminobutyric acid (GABA) degradation after controlling for sex and psychotropic medication usage (Figure 3a, R = 0.439, p = 0.04, p.adj = 0.07).

**Figure 3:**
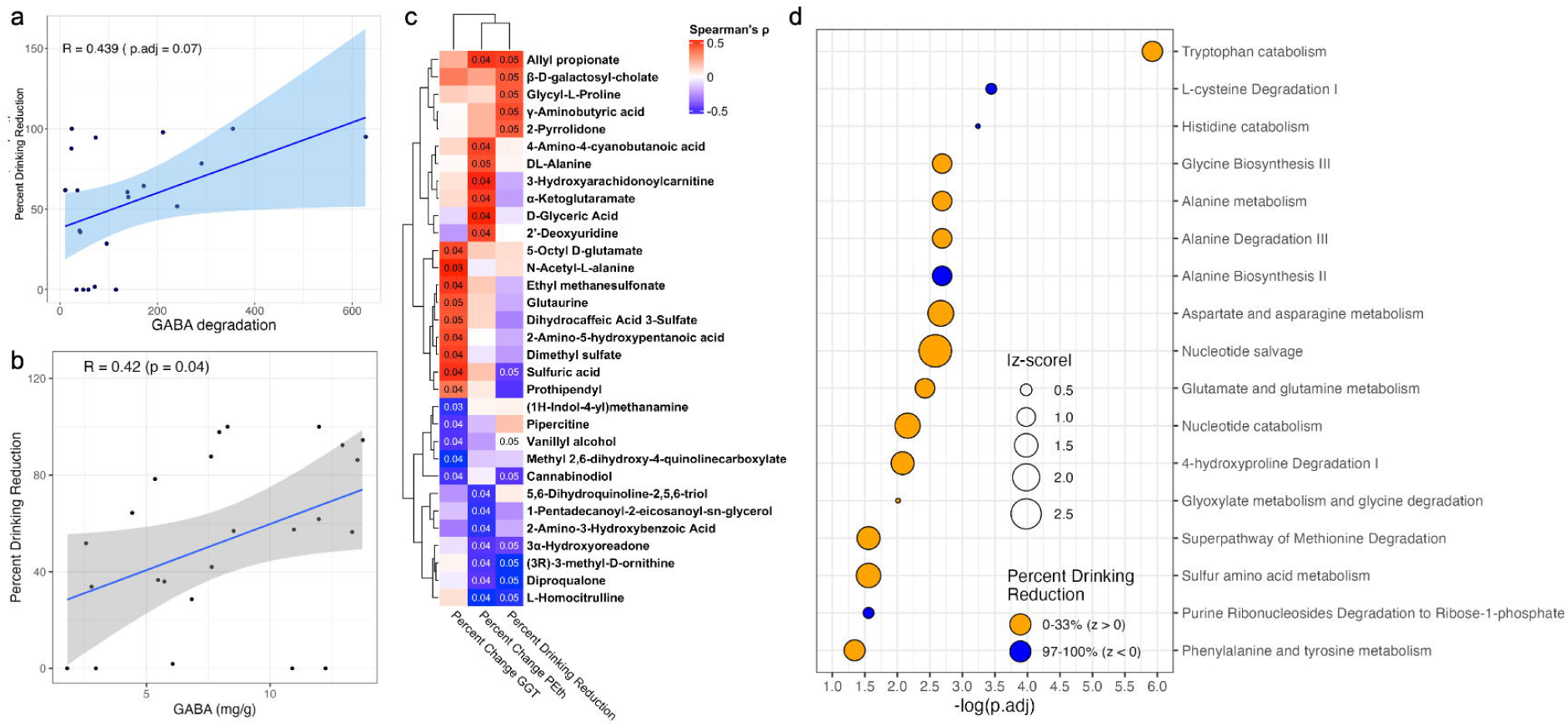
Baseline gut metabolites are associated with percent drinking reduction and change in alcohol-related biomarkers. (a) GABA degradation potential as identified from predictive gut-brain module analysis shows a positive linear relationship with percent drinking reduction controlling for sex and psychotropic medication usage (R = 0.439, p.adj = 0.07). (b) This relationship is recapitulated by direct measurements of stool GABA by an ELISA assay (R = 0.42, p = 0.04). (c) The stool metabolites are correlated with percent drinking reduction, changes in alcohol-related biomarkers PEth and GGT. Colors of the heatmap represent correlation coefficients derived from the multiple linear regression analysis. An adjusted p value is denoted in the cell of the heatmap if the adjusted p value is less than 0.1 for a given correlation analysis. (d) Baseline stool metabolic pathways associated with high- and low-responders. Log2-fold change of metabolites that are significantly different between high- and low-responders were used to construct metabolic pathways using Qiagen IPA software. Dot color indicates the group: a blue dot indicates that the pathway is overrepresented in high responders, orange indicates overrepresentation in low responders. Dot size corresponds to pathway z-score magnitude (*ie* larger dot size corresponds to higher pathway representation).

GABA has been implicated in mediating craving and severity of alcohol use disorder and identified as a potential target for intervention.^2,40–44^ Baseline stool GABA concentration was directly quantified *via* ELISA to confirm the correlation observed from predicted GBM function. Stool GABA concentration was found to be significantly positively correlated with drinking reduction (Figure 3b, R = 0.42, p = 0.04) recapitulating the predicted metagenome functionality.

### Baseline stool metabolome is correlated with alcohol consumption

Given the strong association between baseline stool GABA concentration and drinking reduction, we next analyzed the baseline stool metabolome to determine additional correlations with alcohol consumption.

Baseline stool aliquots from 21 participants were subjected to untargeted metabolomics *via* LC-MS. Annotated stool metabolite levels were used to explore correlations between baseline stool metabolites and alcohol consumption collected as study metadata (Figure 3a). The metabolome was not significantly associated with either sex or psychotropic medication usage. Allyl propionate, β-D-galactosyl cholate, glycyl-L-proline, GABA, and 2-pyrrolidone are all positively correlated with percent drinking reduction with p.adj < 0.05. In addition to the consistent finding of the relationship between GABA and percent drinking reduction (R = 0.470, p.adj = 0.05), 2-pyrrolidone, a biologically relevant cyclization product of GABA,^45^ is also positively correlated with percent drinking reduction (R = 0.434, p.adj = 0.05).

Stratifying the metabolome results based on percent drinking reduction allowed us to determine metabolites that varied between high and low responders. Of the 5,421 annotated metabolites, 1,788 had CV < 20 and corresponding PubChem database entries and were used for subsequent analysis. 63 metabolites were found to differ between high and low responders after adjusting for multiple comparisons (p.adj < 0.05, Kruskal-Wallis). We next performed metabolic pathway analysis using the log2-fold change in low responders with respect to high responders and corresponding p-value after adjusting for multiple comparisons (Kruskal-Wallis) (Figure 3b). Tryptophan catabolism is highly significantly overrepresented in the low responder metabolome (p.adj = 1.2E-6, z-score = 1.069). This is consistent with the predicted metagenome functional analysis that shows microbes with tryptophan degradation functionality are enriched in low responders’ microbiome (Supporting Figure 5a, p.adj = 0.03). Cysteine degradation is highly significantly overrepresented in the high responder metabolome (p.adj = 3.5E-4, z-score = -0.447) whereas cysteine synthesis is overrepresented in the low responder metabolome (p.adj = 0.1, z-score = 2). Similarly, alanine synthesis is overrepresented in the high responder metabolome (p.adj = 0.002, z-score = -1) while alanine metabolism and degradation are overrepresented in the low responder metabolome (p.adj = 0.002, z-score = 1). The overall metabolome is not significantly different between high and low responders (Supporting Figure 5b).^31^

## Discussion

In light of the recent interest in gut microbiome as both a diagnostic tool as well as a potential avenue for treatment of AUD, we investigated the novel connection between baseline gut microbiome composition and gut metabolites in a zonisamide clinical trial. Across all participants, regardless of treatment, percent drinking reduction was strongly correlated with baseline (*ie* before treatment) microbiome composition and functionality. This difference appears to be driven by composition as opposed to microbial richness, as alpha diversity is relatively consistent across participants, but beta diversity differs widely. Individuals with AUD are often reported to have lower alpha diversity as compared with healthy controls^46,47^ but this is not always the case.^17,27^ There is even less consensus on the relationship between AUD severity and alpha diversity.

The variation in drinking reduction across patients from both placebo and treatment groups was binned into tertiles to compare the highest and lowest responding participant microbiomes. There is a significant difference in microbiota composition between patients that had a high response to the intervention (67-100% reduction in drinking) and those that had a low response (0-33% reduction in drinking. At a genus level, low responders had a higher relative abundance of *Sutterella* and *Lachnoclostridium*. Genera positively correlated with drinking reduction in this study (*Collinsella*, *Bilophila*, and *Eubacterium*) have been similarly found to be enriched in patients with AUD relative to healthy controls while *Akkermansia* was depleted relative to healthy controls.^46,47^ In this study, we found that *Akkermansia* was negatively correlated with percent drinking reduction suggesting that a lower abundance of *Akkermansia* may be beneficial for reduced alcohol consumption.

Leclercq *et al* found a positive correlation between *Akkermansia* and quinolinic acid, a neurotoxic tryptophan metabolite elevated during alcohol withdrawal.^48^ Tryptophan metabolism has been implicated in systemic inflammation, depression, and craving in AUD.^48–50^ The potential involvement of *Akkermansia* in the overrepresentation of tryptophan metabolism and the negative correlation between *Akkermansia* relative abundance and percent drinking reduction further suggest that *Akkermansia* may be detrimental in the heavy drinking. While *Akkermansia* has most often been discussed as a beneficial microbe and target for probiotic supplementation,^51–53^ recent study has indicated that *Akkermansia* may be detrimental in the context of neuropsychiatric disorders.^54,55^ The relationship between *Akkermansia* abundance, tryptophan metabolism, and alcohol consumption is an area of potential future study.

The ideal composition of the gut microbiome is very difficult to conceptualize given the highly individual and fluctuating nature of the gut microbiome. However, microbial metabolism of the collective gut microbiome is relatively conserved due to multiplicity of function and can lend insight into the various metabolic niches that individual microbes may occupy. GABA is one such metabolite that is produced by the gut microbiome and is also vital in the context of AUD. Microbial GABA production has an unclear correlation to brain GABA concentrations.^56–58^ Existing evidence suggests that the microbial GABA production may contribute to circulating GABA levels, though it is believed that GABA itself does not cross the blood brain barrier. Microbial GABA more likely indirectly influences the brain through the vagus nerve without entering circulation. The GABA present in the gut may also arise from dietary sources, but dietary GABA is relatively low. GABA remains a strong target for AUD interventions.

Several medications that act directly or indirectly on GABA or glutamate receptors have been approved for treatment of AUD.^2,40,43,59^ Zonisamide itself is a GABAergic medication typically used in treating epilepsy.^60^ In our study, we consistently identified the positive correlation between GABA and drinking reduction using three independent approaches, suggesting importance of gut-derived GABA in association with alcohol consumption. Looking at the baseline microbiome production of GABA before starting either placebo or a GABAergic treatment can provide valuable information.

Sex and psychotropic medication usage are significantly correlated with the composition of the baseline microbiome. Sex-linked dimorphism in gut microbiome composition is well-established in both human and animal models.^61–67^ The male microbiome typically has an increased relative abundance of *Bacteroidetes* when comparing among healthy weight individuals,^65^ but Haro *et al* found a reversal of this trend with increasing BMI.^63^ Similarly, Dong *et al* found a significant increase in *Bacteroides* and *Blautia* abundance in women with BMI ≥ 25 as compared with those under BMI 25.^68^ Given that the cohort in this study has a median BMI of 28.7, it is vital to consider the effects of western diet and obesity on the expected sex-linked gut microbiome composition. Additionally, non-antibiotic medications are known to alter the gut microbiome, though there is not yet a clear consensus on the shifts caused by medication. ^69–75^ The correlations found in this study suggest that a microbiome closer to that found in healthy controls might not necessarily be reflective of a positive intervention outcome. Our results suggest that regardless of other contributing factors such as sex, medication usage, or study treatment (placebo or zonisamide) baseline microbiome may play an important role in intervention outcome at an individual level.

This study presents several limitations. Small sample size is a consistent issue in microbiome-related work given the large intra-individual variation in microbiome composition. Further, analysis of fecal metabolites without measurement of circulating metabolite concentrations limits the ability to draw conclusions between microbiome function and host processes. Future studies with large sample size from multiple clinical sites are warranted to verify the relationship between baseline microbiome and alcohol consumption. Lastly, the study is an association study, and we cannot conclude a causal relationship between the baseline microbiome and alcohol drinking reduction. Animal studies such as fecal microbiota transplantation will allow to gain mechanistic understanding on contribution of the baseline microbiome in reducing alcohol consumption.

In conclusion, our study identified important associations between baseline gut microbiome and gut-derived GABA with alcohol consumption reduction in a clinical trial. Screening baseline microbiome composition and metabolites may hold significant value as a predictive tool in clinical settings to better personalize intervention and improve reduction in alcohol consumption, durability of behavioral changes, and ultimately patient outcome.

## Methods

### Human Trial

Patients were recruited from the community at three sites (two in Connecticut and one in Virginia) as part of a double-blind, randomized, placebo-controlled study investigating the use of zonisamide in reducing drinking (ClinicalTrials.gov identifier: NCT02900352). Inclusion criteria limited patients to ages 21-70 who had regular heavy drinking, a current DSM-5 diagnosis of alcohol use disorder, and a desire to reduce or stop drinking. Potential patients who were currently lactating or with clinically significant physical disease, seizure disorder, use of any medications that could affect drinking or cause harm, schizophrenia, bipolar disorder, substantial risk of suicide or violence, opioid or benzodiazepine dependence were excluded. Women of child-bearing age were required to practice a reliable method of birth control. Patients gave written consent to participate in the study and were financially compensated. Patients were randomized into treatment and control groups matching for sex and current psychotropic drug usage. The treatment group received flexibly titrated zonisamide over 7 weekly visits starting at 100 mg daily and increasing over the 8 weeks to a 500 mg daily maximum/200 mg daily minimum for the remaining 8 weeks of the study. Medical management^76^ served as a psychosocial intervention platform. Timeline Follow-back Method^77^ was used to measure self-reported drinking including number of drinking days during the 90-day pretreatment period and at each visit. Non-fasting serum PEth and GGT was measured at baseline, midpoint, and endpoint visits to validate self-reported drinking.

Fisher’s exact test was used to test the difference in demographic and clinical categorical variables between zonisamide and placebo groups (Table 1). Kruskal-Wallis rank sum test was used for testing continuous demographic and clinical variables. Percent drinking reduction was quantified as the difference between self-reported drinks per week at baseline and average drinks per week over the last four weeks of the study as calculated at study endpoint divided by the baseline self-reported drinks per week.

### 16S rRNA Sequencing of Stool Samples

Stools were collected by participants at UConn Health site and stored on ice up to 24 h prior to baseline and 16-week (endpoint) visits. Stool aliquots were prepared upon receipt at the clinical research center at UConn Health and stored at -80[ until the time of analysis. Microbial DNA was isolated from stool samples using the PowerSoil DNA Isolation kit (Qiagen) following manufacturer’s instructions. Bacterial 16S ribosomal RNA (rRNA) gene sequencing was performed on V4 hypervariable regions using 515F (5’-GTGYCAGCMGCCGCGGTAA-3’) and 806R (5’-GGACTACNVGGGTWTCTAAT-3’) primers to prepare an amplicon library that was purified using Zymo Select-a-Size MagBeads (Zymo), quantified (Qubit 2.0 fluorimeter, Invitrogen), and pooled with equal masses added from each sample. Two additional cleanup steps were performed on the initial pool again using Zymo Select-a-Size MagBeads (Zymo). The pooled and purified library was sequenced on the Illumina MiSeq platform (Illumina) using 2 × 250 bp, 500 cycles kits.

### 16S rRNA Data Processing and Analysis

Raw 16S rRNA sequencing reads were initially processed by bcl2fastq2 (v2.20) and RTA (v1.18.54.4) software (Illumina). Demultiplexed fastQ files were imported into the QIIME2 pipeline (version 2022.11).^78,79^ The DADA2^80^ plugin was used to denoise reads and remove chimeras using the consensus method. Forward and reverse reads were truncated at position 250. All other parameters were set to default. Samples were rarified to a sampling depth of 8800 reads/sample prior to alpha and beta diversity analyses. The phylogeny was inferred using the align-to-tree-mafft-fasttree pipeline in QIIME2. Taxonomy was assigned with pre-trained naïve Bayesian classifier based on the SILVA reference database V138.1 using the q2-feature-classifier plugin with a 0.5 confidence value cut-off.

Subsequent analysis of 16S rRNA sequencing data was done in R (version 4.3.1) using RStudio interface (version 2023.06.1).^81^ and Qiime2R^82^ and phyloseq^83^ packages. ASV counts were aggregated at various taxonomic levels (*ie* genus-level) and converted to relative abundance using the phyloseq^83^ and MicroViz^84^ packages. PERMANOVA (permutational multivariate analysis of variance) was performed using the Adonis function in vegan package^85^ to evaluate differences in beta diversity across metadata variables. Principal coordinate analysis and visualization with 90% confidence intervals (stat_ellipse, ggplot2^86^) were generated using microViz,^84^ ggplot2,^86^ and tidyverse^87^ packages in R. ^77^ ^78^ Correlations between genus-level relative abundance and metadata variables were tested using multiple linear regression controlling for sex and psychotropic medication usage and adjusting p-values for multiple comparisons using false discovery rate. An adjusted p-value <0.1 was considered statistically significant. Linear discriminant analysis Effect Size (LEfSe)^36^ was performed using the corresponding galaxy module with a significance cutoff of p-value <0.05.

### Gut-Brain Module Analysis Based on 16S Data

The ASVs for samples of interest was exported from R and used for subsequent phylogenetic investigation of communities by reconstruction of unobserved states (PICRUSt) using PICRUSt2.^38,39^ The PICRUSt2 pipeline was run using picrust2_pipeline.py and add_descriptions.py. The resultant unstratified KO metagenome predictions and their associated descriptions was subsequently used for predictive functional analysis *via* gut-brain modules.^56^ Correlations between number of hits in each module and metadata variables were tested using Spearman’s rank correlation with a significance cutoff of p-value <0.05 after adjusting for sex and psychotropic medication usage and controlling for multiple comparisons. Wilcoxon rank-sum tests were performed to compare the differences number of hits in each module between high-responder (67-100% drinking reduction) and low-responder (0-33% drinking reduction) patients with a significance cutoff of p-value <0.05 after adjusting for sex and psychotropic medication usage and controlling for multiple comparisons.

### Untargeted LC-MS Analysis of Stool Metabolome

∼20 mg aliquots of stool samples from 21 participants that provided both baseline and endpoint stools were subjected to untargeted LC-MS metabolomic analysis. Stool samples were homogenized in homogenization buffer (80% methanol in PBS with 1.8105 mM ^13^C_3_-lactate and 142 μM ^13^C_5_-glutamic acid) prior to the addition of 800 μL homogenization buffer. Homogenized samples were incubated 30 min at -20[ and subsequently sonicated for 30 min on ice. Debris was pelleted *via* centrifugation and 800 μL supernatant was dried under vacuum (CentriVap Concentrator, Labconco). The dried residue was suspended in 150 μL 40% PBS/60% acetonitrile. A quality control sample was pooled from all study samples.

The untargeted LC-MS metabolomic method was adapted from previously published methods.^88–92^ In summary, each sample was injected twice (10 μL for negative ionization mode, 4 μL for positive ionization mode) onto an XBridge BEH Amide column (150 x 2.1 mm, 2.5 µm particle size, Waters) maintained at 40[. Samples were maintained in an autosampler at 4[. Mobile phase flow rate was 0.3 mL/min and was composed of MP A (5% acetonitrile in water, 10 mM ammonium acetate and ammonium hydroxide) and MP B (95% acetonitrile in water, 10 mM ammonium acetate and ammonium hydroxide). The mobile phase gradient is as follows: 1 min isocratic elution, 90% MP B; 10 min ramp to 40% MP B; 4 min hold at 40% MP B; ramp to 90% MP B prior to next injection. Untargeted data was collected from 70 to 1050 m/z using Thermo Vanquish UPLC-Exploris 240 Orbitrap MS instrument (Thermo Scientific) equipped with an electrospray ionization source.

Data were processed using Thermo Compound Discover 3.3 software (Thermo Scientific) for peak picking, alignment, and normalization. Only peaks with CV <20% across quality control pools appearing in >80% of all samples were included in all subsequent analysis. Identifications and annotations used available data for retention time, exact mass, and fragmentation & isotopic patterns. Data extraction absolute intensity threshold was 1,000 and mass accuracy limit was 5 ppm. Peaks in the obtained MS spectra were annotated using an extensive in-house library of ∼600 aqueous metabolites in addition to the HMDB library, LIPID MAPS database,^93,94^ METLIN database,^95–97^ ChemSpider database^98^ and commercial databases (mzCloud (HighChem LLC), Metabolika (Thermo Scientific)). Annotated metabolites were used for downstream analysis in R, MetaboAnalyst,^99^ and Ingenuity Pathway Analysis (Qiagen). Correlations between normalized peak intensity and metadata variables were tested using Spearman’s rank correlation coefficient. P-values were corrected for multiple comparisons using false discovery rate. A subset of annotated metabolites with CV > 20 that were accessible in the PubChem database (n = 1789) were used for IPA analysis. Log2-fold change of metabolites in low-responders with respect to high-responders and corresponding p-values (Kruskal-Wallis) were provided to Qiagen IPA software.

### Stool GABA Quantitation

∼100 mg aliquots of stool samples from 31 participants that completed the study were analyzed for GABA content using ELISA kit (LDN, Nordhorn, Germany) following manufacturer’s instructions. Stool was thawed and homogenized in 300 μL lysis solution (0.01N HCl, 1 mM EDTA, & 4 mM sodium metabisulfite) (Thermo Fisher Scientific). Homogenized fecal slurry was clarified by centrifugation at 5000g for 10 min at 4[ prior to subsequent use. In brief, clarified fecal slurry and standards (provided by LDN) were extracted, derivatized, and incubated with antiserum. Derivatized samples and standards were subjected to a quantitative ELISA read at 450 nm in a 96-well plate reader (iMark, Biorad). Absorbance of derivatized standards was used to generate a standard curve that was used to quantify the experimental samples. Correlation between percent drinking reduction and stool GABA content was tested using Spearman’s rank correlation coefficient.

## Data Availability Statement

Raw sequencing reads from the 16S amplicon sequencing are available via the National Center for Biotechnology Information (NCBI) Short Read Archive (SRA) under accession #PRJNA1065830.

## Author Contributions

AJA, JMC, and YZ were responsible for study design. LRD, HY, JC, and HG performed data analysis. LRD interpreted findings and drafted the manuscript. All authors critically reviewed the manuscript and agree to publication of the final version.

## Acknowledgments

This work was supported by the National Institute on Alcohol Abuse and Alcoholism (NIAAA) under grants AA027858, AA12722313, AA007290, and AA027055. We thank Pam Fall and Judy Kalinowski at the UConn Health Clinical Research Center for stool and serum sample management.

## Funding statement

This work was supported by the National Institute on Alcohol Abuse and Alcoholism (NIAAA) under grants AA027858, AA12722313, AA007290, and AA027055.

## Conflict of interest disclosure

AJA is a consultant to Sobrera Pharmaceuticals. All other authors report no conflict of interest.

## Clinical trial registration

Patients were recruited from the community at three sites (two in Connecticut and one in Virginia) as part of a double-blind, randomized, placebo-controlled study investigating the use of zonisamide in reducing drinking (ClinicalTrials.gov identifier: NCT02900352).

## Supplementary Figures

**Supporting Figure 1:**
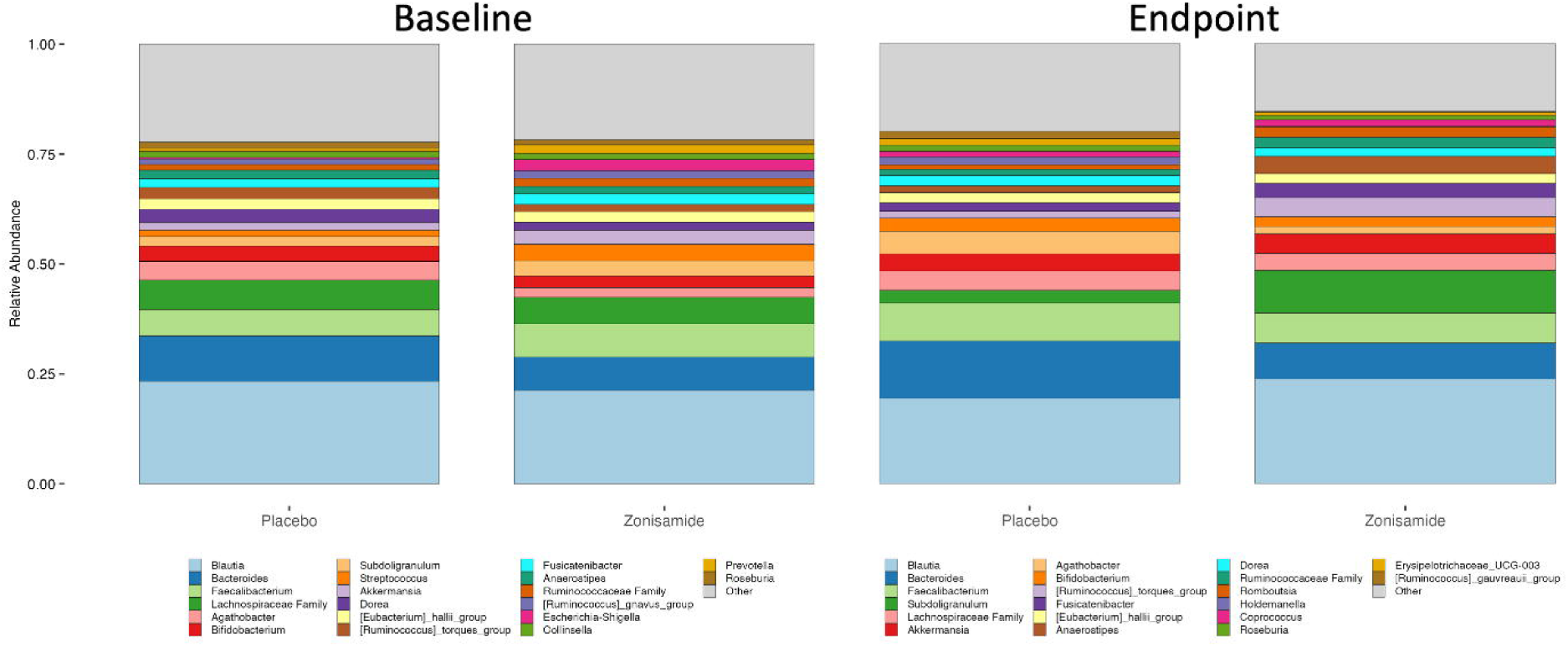
The averaged relative abundance of the gut microbiome at the genus level in zonisamide and placebo groups at baseline and end point.

**Supporting Figure 2:**
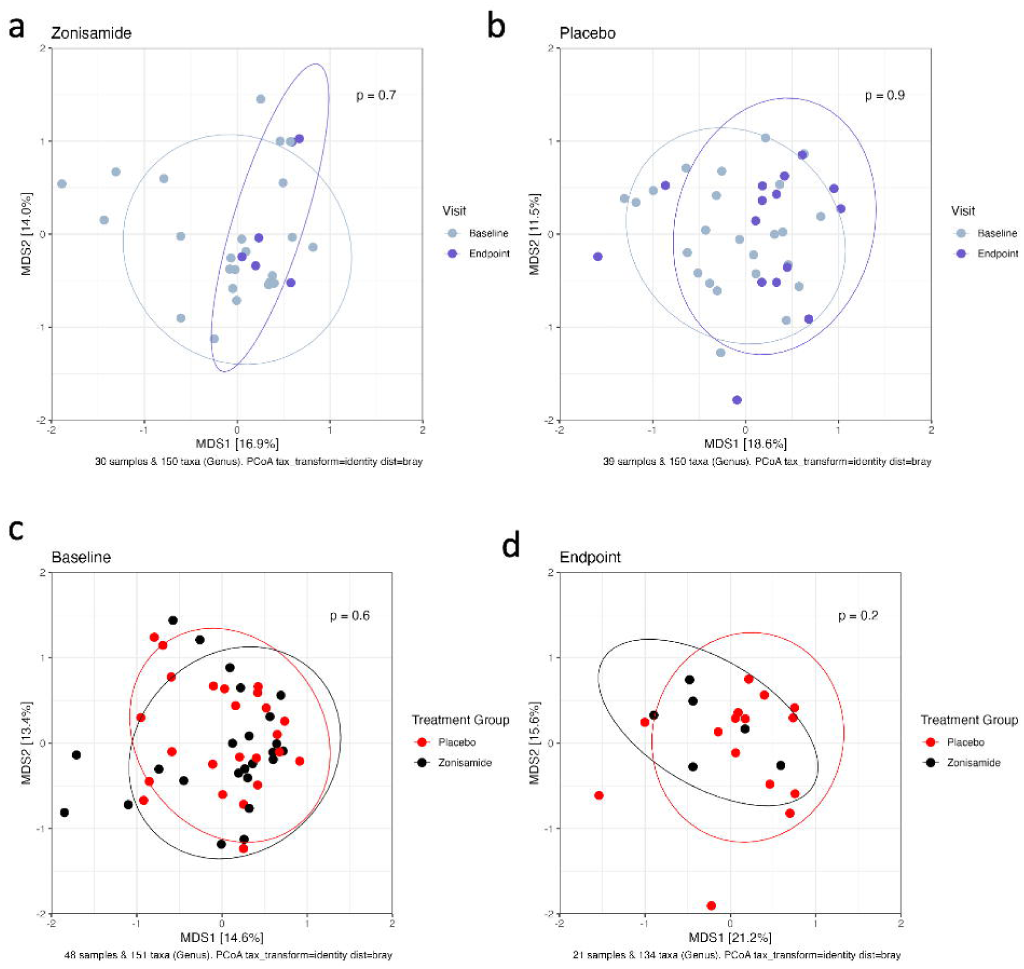
(a-b) PCoA plots and PERMANOVA analysis indicate the overall gut microbiome community structure of participants remain the same before and after treatment in the two groups. (c-d) PCoA plots and PERMANOVA analysis indicate the overall gut microbiome community structure of participants of the two groups is similar at baseline or at end point.

**Supporting Figure 3:**
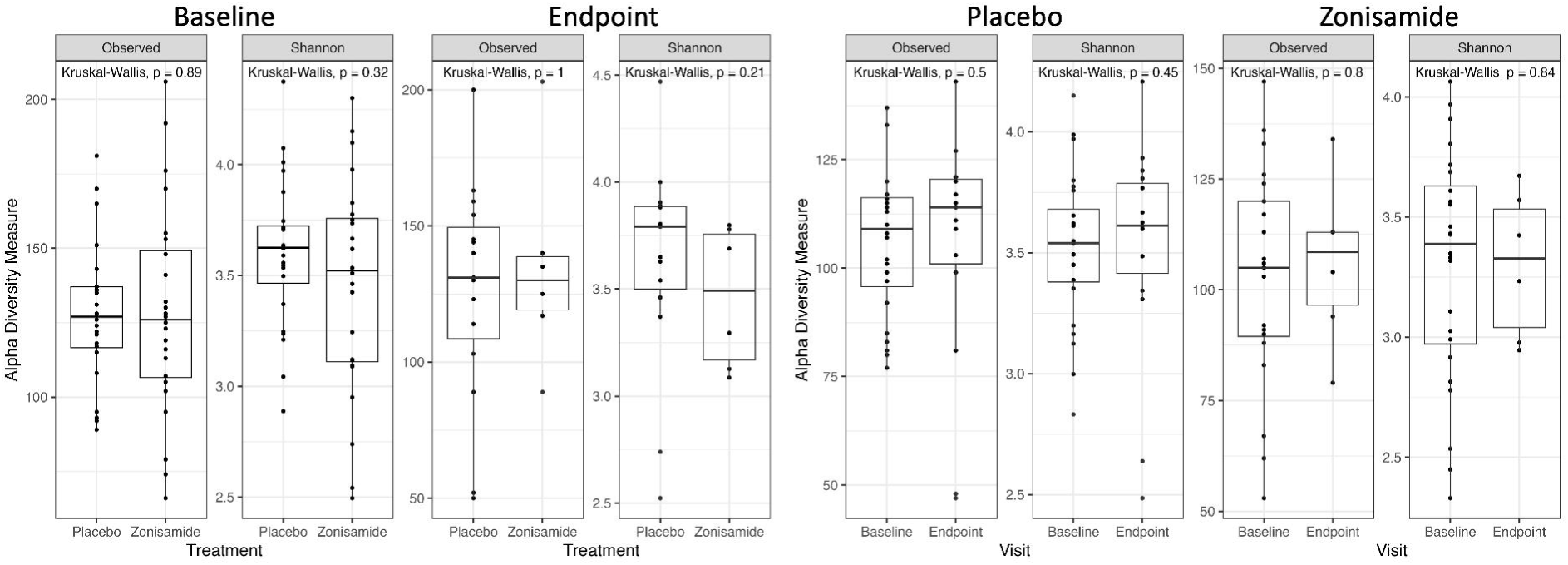
Alpha diversity measures as described by observed OTUs and Shannon diversity do not differ between treatment groups at baseline and endpoint visits.

**Supporting Figure 4:**
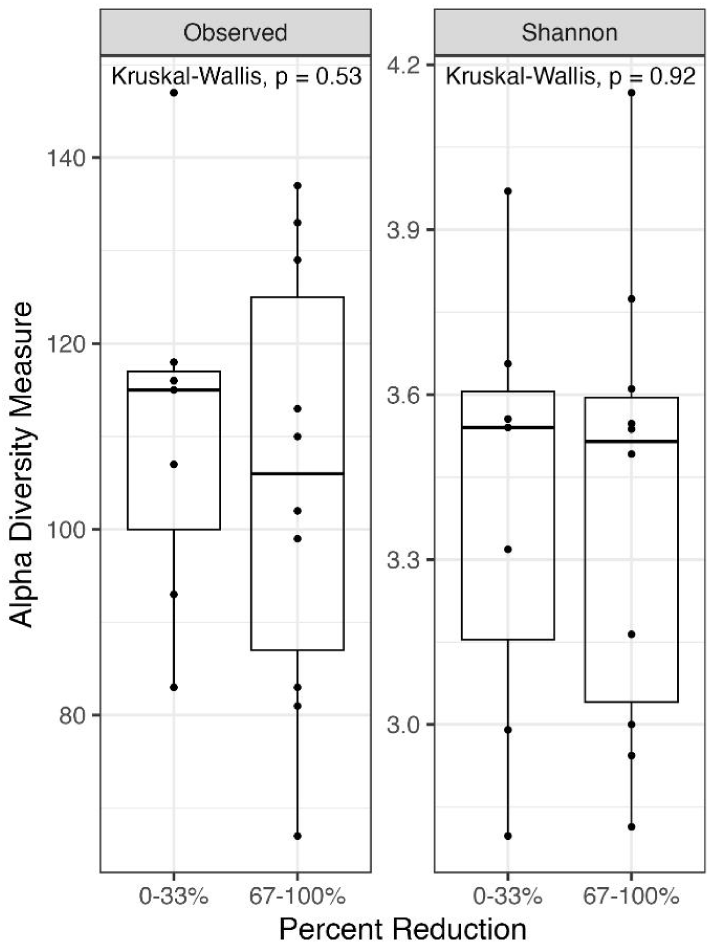
Alpha diversity measures as described by observed OTUs and Shannon diversity do not differ between high-(67-100% drinking reduction) and low-responder (0-33% drinking reduction) groups.

**Supporting Figure 5:**
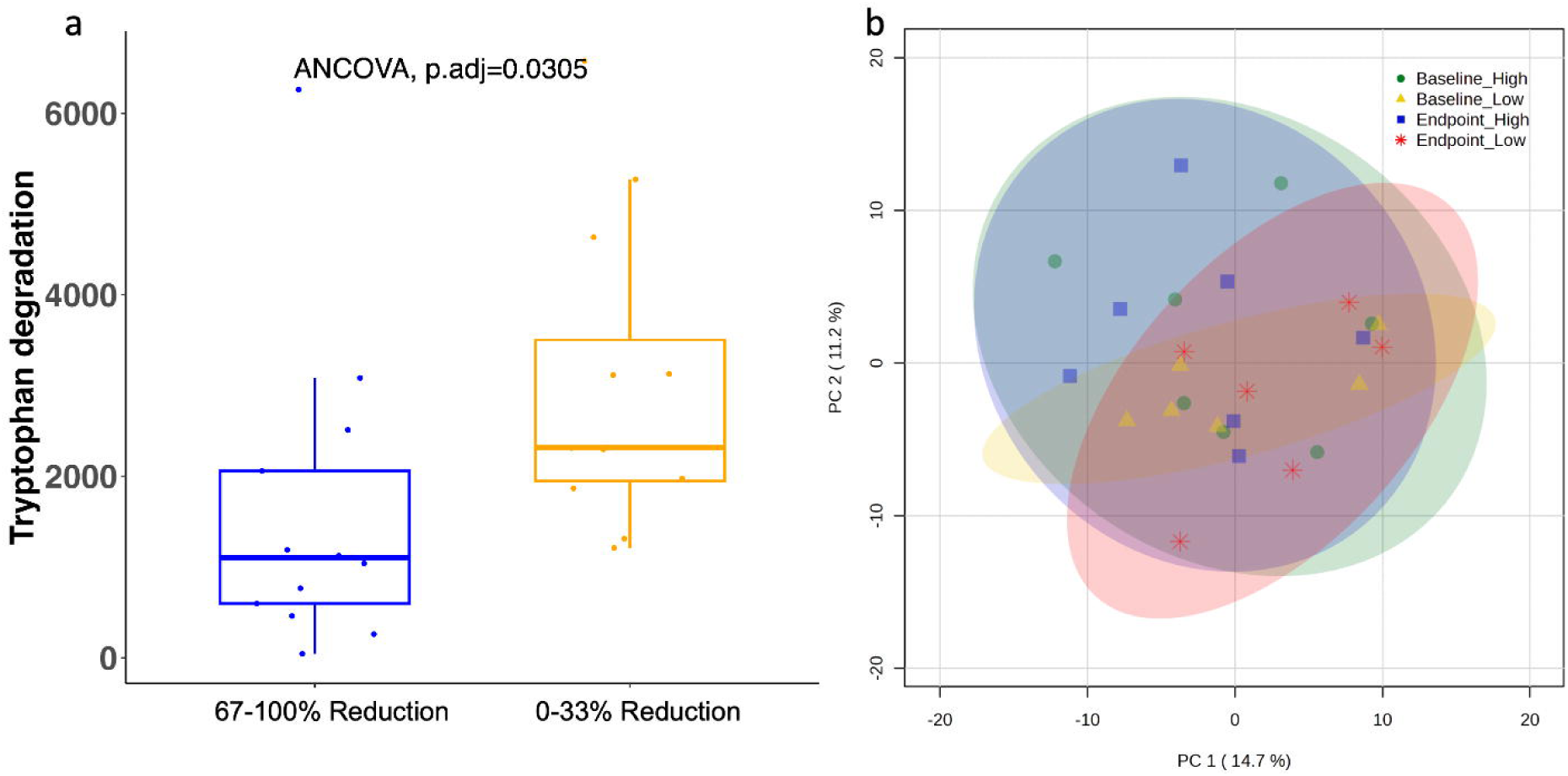
(a) Tryptophan degradation identified by the gut-brain module analysis is overrepresented in the low-responder group at both baseline and endpoint controlling for sex and psychotropic medication usage (p.adj = 0.03). (b) The overall stool metabolome does not vary between high- and low-responder groups at baseline and endpoint.

## Notes

### Clinical Trial

NCT02900352

### Author Declarations

Ethics committee/IRB of VIRGINIA COMMONWEALTH UNIVERSITY and Yale University gave ethical approval for this work

